# The Predictive and Prognostic Value of T- and B-cell Transcriptomic Signatures for Clinical Response to Immune Checkpoint Blockade in Pleural Mesothelioma

**DOI:** 10.1101/2025.05.12.25326806

**Authors:** Jasper van Genugten, Daniel Faulkner, Jens C. Hahne, Maria Disselhorst, Lodewyk Wessels, Dean Fennell, Paul Baas

## Abstract

**Background:** Malignant pleural mesothelioma (PM) is an aggressive cancer with limited treatment options. Although immune checkpoint blockade (ICB) with nivolumab plus ipilimumab improves overall survival and has become standard-of-care, responses are variable and there are no predictive biomarkers for treatment outcome among PM patients with epithelioid histology.

**Methods:** We developed an eight-gene T-cell and six-gene B-cell transcriptomic signature. To quantify T- and B-cell infiltration, a geometric-mean T- and B-cell expression score was calculated for each patient. Immune profiles were generated in three ICB-naïve PM cohorts (n = 448), and Kaplan-Meier analysis was used to evaluate prognostic value of the signatures. Predictive value was tested in five independent ICB-treated cohorts (n = 104).

**Results:** In ICB-naïve patients, high B-cell infiltration was associated with longer overall survival (hazard ratio (HR): 0.50, 95% confidence interval (CI): 0.31-0.82), whereas T-cell infiltration had no prognostic value. Among patients treated with 2^nd^ line treatment with nivolumab plus ipilimumab, high T-cell infiltration predicted better objective response and improved overall survival (HR: 0.06, 95% CI: 0.01-0.36). This effect was absent in patients treated with 2^nd^ line nivolumab alone or any other anti-PD1/PDL1 drug combined with non-immunotherapeutics. B-cell infiltration showed no predictive value in any ICB-treated group.

**Conclusions:** The eight-gene T-cell signature is a specific predictor of outcome after 2^nd^ line treatment with nivolumab plus ipilimumab, while B-cell infiltration is prognostic in ICB-naïve disease.

## Introduction

Malignant pleural mesothelioma (PM) is an asbestos-related pleural cancer with a 5-year overall survival between 5-12%^1-3^. Platinum-based chemotherapy has historically been standard-of-care^4^, while the role of surgery remains controversial^5-6^. The CheckMate 743 phase III trial recently demonstrated that immune checkpoint blockade (ICB) treatment with nivolumab plus ipilimumab improved survival for some PM patients with a median overall survival (mOS) benefit of 18.1 months compared to 14.1 months with chemotherapy^7^. However, only ∼42% of patients respond, and immune-related toxicity combined with high treatment cost highlight the need for robust predictive biomarkers to stratify patients^7-8^.

PD-L1 expression is the most studied predictive biomarker for ICB in PM^7,9-16^, and might have limited value in predicting response and survival after treatment with nivolumab plus ipilimumab^7,14-15^. In the CheckMate 743 trial, PD-L1 ≥ 1% predicted a benefit of ICB treatment compared to chemotherapy^7^. However, within the nivolumab plus ipilimumab arm PD-L1 expression failed to predict differences in treatment response or overall survival (17.3 vs. 18.0 months)^7^. Given the mixed treatment responses and limited predictive value of PD-L1 expression, novel biomarkers to stratify patients for nivolumab plus ipilimumab are urgently needed.

Proposed alternative biomarkers include tumor mutational burden (TMB), DNA-repair defects, and chromosomal rearrangements^17-18^. However, while genetic alterations and TMB are predictive in some cancer types^19-20^, the low mutation rate and lack of microsatellite instability of PM limits their utility in this disease^21^. The tumor immune microenvironment may hold greater promise in PM^22-23^, as in other cancer types T- and B-cell infiltration were shown to predict ICB benefit^24-28^.

Here, we establish transcriptomic signatures for T- and B-cells to characterize T- and B-cell infiltration in bulk RNA-sequencing data from three independent ICB-naïve^21,29-30^ and five ICB-treated PM cohorts^15-16,31-33^. We retrospectively evaluate their prognostic value in ICB-untreated patients and their predictive value for response to 2^nd^ line ICB treatment in PM patients.

## Materials & Methods

### Patient cohorts

All patients were diagnosed with histologically confirmed PM. Baseline characteristics, including age, sex, asbestos exposure, clinical stage (AJCC), histology, ECOG performance score, and prior chemotherapy, are summarized in **Table 1**. As expected, most patients were male and had epithelioid histology.

**Table 1:**
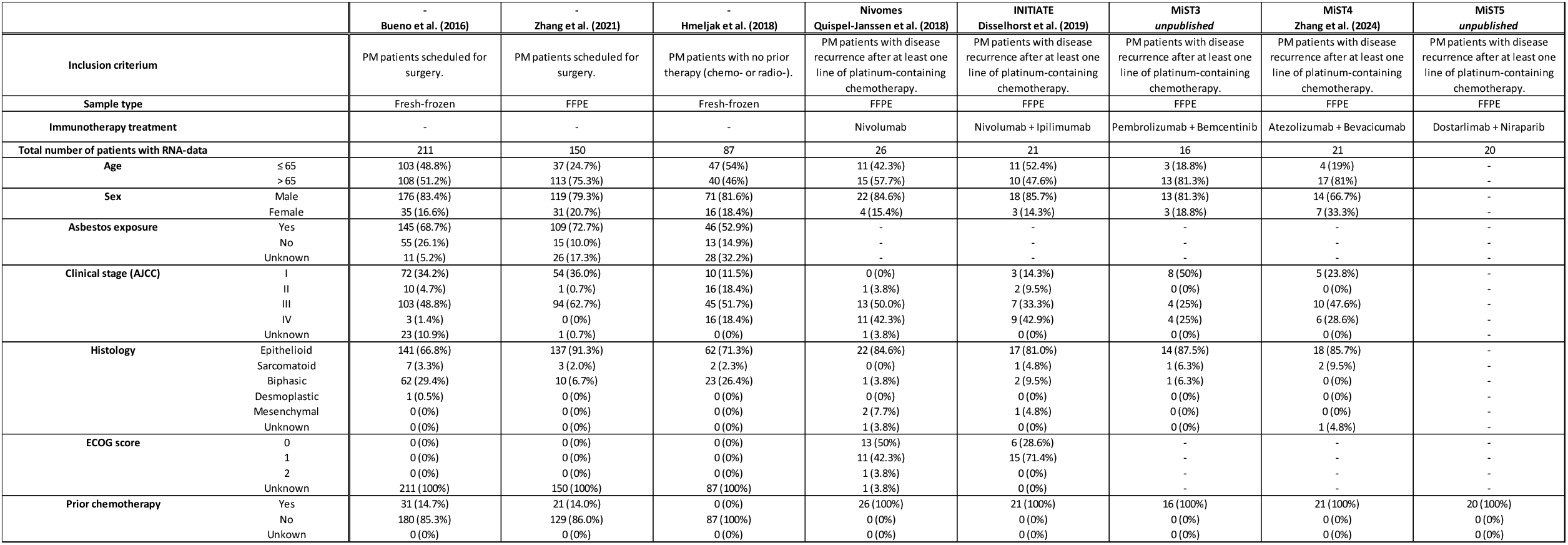
Baseline characteristics of the PM cohorts.

The T- and B-cell signatures were analyzed in bulk RNA-sequencing data from three ICB-naïve cohorts: Bueno (n = 211)^21^, Zhang (n = 150)^29^, and Hmeljak (n = 87)^30^. We also studied RNA-sequencing data from five phase II clinical trial cohorts: NivoMes (2^nd^ line nivolumab; n = 26)^16^, INITIATE (2^nd^ line nivolumab plus ipilimumab; n = 21)^15^, MiST3 (2^nd^ line pembrolizumab plus bemcentinib; n = 16)^31^, MiST4 (2^nd^ line atezolizumab plus bevacizumab; n = 21)^32^, and MiST5 (2^nd^ line dostarlimab plus niraparib; n = 20)^33^. All analyses were performed in diagnostic or pre-treatment biopsies. A subset of patients (14.7% and 14.0%) from the Bueno^21^ and Zhang^29^ cohorts received chemotherapy prior to biopsy collection, and all patients in these two cohorts subsequently underwent surgery. All patients in the clinical trial cohorts received at least one line of platinum-containing chemotherapy before ICB-treatment.

### Ethics approval

This study was approved by the Netherlands Cancer Institute IRB (IRBd24-234).

### RNA-sequencing data processing

Read count RNA-sequencing data from Bueno^21^, Zhang^29^, NivoMes^16^, INITIATE^15^, and the MiST3/4/5^31-33^ cohorts were normalized using the DESeq2 R package^34^. For the Hmeljak cohort^30^, RSEM-normalized data were retrieved from the Firebrowse portal. Since our analyses of T- and B-cell infiltration relied on ranking of samples based on T- and B-cell signature scores within each cohort, we do not expect these normalization differences (DESeq2 vs. RSEM) to impact the results of these analyses. Each cohort was normalized and analyzed independently, since no cross-cohort pooling was performed, inter-dataset correction was not required.

### Transcriptomic T- and B-cell signatures

We developed transcriptomic signatures to quantify T- and B-cell infiltration (**Table 2**). The T-cell signature was based on T-cell receptor (CD3D, CD3E, CD3G, CD8A, and CD8B) and effector genes (GZMA, GZMB, PRF1)^35^, while the B-cell signature was based on B-cell receptor (CD19, BANK1, FCRL1) and membrane genes (CR2, MS4A1, CXCR5)^36^. Single-cell RNA-sequencing data from the Human Protein Atlas^37^ was used to validate cell-type specificity of these markers.

**Table 2:** Composition of the T- and B-cell signatures.

| T-cell signature |  |  |
| --- | --- | --- |
| Gene | Gene name | Function |
| CD3D | CD3 Delta Subunit Of T-Cell Receptor Complex | T-cell receptor (TCR)/CD3 complex |
| CD3E | CD3 Epsilon Subunit Of T-Cell Receptor Complex | T-cell receptor (TCR)/CD3 complex |
| CD3G | CD3 Gamma Subunit Of T-Cell Receptor Complex | T-cell receptor (TCR)/CD3 complex |
| CD8A | CD8 Subunit Alpha | T-cell receptor (TCR)/CD3 complex |
| CD8B | CD8 Subunit Beta | T-cell receptor (TCR)/CD3 complex |
| GZMA | Granzyme A | Cytolytic enzyme |
| GZMB | Granzyme B | Cytolytic enzyme |
| PRF1 | Perforin 1 | Cytolytic pore-forming protein |

| B-cell signature |  |  |
| --- | --- | --- |
| Gene | Gene name | Function |
| CD19 | CD19 Molecule | B-cell receptor (BCR) complex |
| BANK1 | B Cell Scaffold Protein With Ankyrin Repeats 1 | B-cell receptor (BCR) complex |
| FCRL1 | Fc Receptor Like 1 | B-cell receptor (BCR) complex |
| CR2 (CD21) | Complement C3d Receptor 2 | B-cell-specific membrane proteins |
| MS4A1 (CD20) | Membrane Spanning 4-Domains A1 | B-cell-specific membrane proteins |
| CXCR5 | C-X-C Motif Chemokine Receptor 5 | B-cell-specific membrane proteins |

### Quantification of T- and B-cell infiltration

For each sample a T- and B-cell score was calculated based on the log-transformed mean of signature gene expression, with a pseudo count of 1 added to each value^38^:

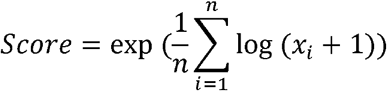

Spearman correlations were used to assess the correlation between T-cell score and CD4, CD8, and PD-L1 immunohistochemistry (IHC) staining in the INITIATE cohort^15^, and to assess the correlation between T- and B-cell scores and immune checkpoint (PD1, PDL1, CTLA4) expression in the Bueno cohort^21^.

### Clustering of ICB-naïve PM patients

We profiled the landscape of T- and B-cells in ICB-naïve patients through unsupervised hierarchical clustering of the samples based on the T- and B-cell signature gene panels. The ConsensusClusterPlus R package^39^ was used for clustering (parameters: *maxK*=20, *reps*=2000, *pItem*=0.8, *pFeature*=1, *clusterAlg*=“hc”, *distance*=“pearson”), and optimal cluster number (*k*) was determined based on the delta area plots. Heatmaps were created for each cohort at optimal *k* using the log_10_-transformed gene expression values, and plotted in Microsoft Excel with colors indicating the range between lowest and highest expression values for each gene separately. Patients were classified into one of three recurrent immune phenotypes, group one: T-cell ^--^;B-cell ^--^, group two: T-cell ^+^;B-cell ^++^, group three: T-cell ^++^;B-cell ^+^, with all other intermediate samples classified as “rest”.

### Survival analyses

To test the associations between T- and B-cell infiltration and overall survival in PM patients, we used univariate Kaplan-Meier survival analysis with log-rank tests to compare survival differences. Hazard ratios (HRs) and 95% confidence intervals (CIs) were calculated for group comparisons and to create Forest plots. Based on available data and numbers, patients were ranked by T- and B-cell scores and classified into three subgroups for T- and B-cell scores:

1. Top quartile (25%; high) versus (vs.) bottom three quartiles (75%; low).
2. Top half (50%; high) vs. bottom half (50%; low).
3. Top three quartiles (75%; high) vs. bottom quartile (25%; low).

Statistical significance for group comparisons was adjusted for multiple-testing by Bonferroni correction (6 tests per cohort). A threshold for T-cell score of the top three quartiles (75% high) vs. bottom quartile (25% low) was determined based on analysis from the INITIATE^15^ and NivoMes^16^ cohorts. Multivariate Cox-analysis was performed to assess the interaction between T- and B-cell score and histology in the Bueno^21^, Zhang^29^ cohorts.

### Statistical considerations

Statistical significance was set at p < 0.05. All RNA-sequencing data processing and normalization, and T- and B-cell score calculations were performed in R-Studio. GraphPad Prism v.10.0.0 was used for statistics and plotting, and heatmaps for T- and B-cell-based subgroups of ICB-naïve patients were created in Microsoft Excel for Windows.

## Results

### Validation of T- and B-cell signatures

We developed an eight-gene T-cell and six-gene B-cell transcriptomic signature to evaluate immune infiltration in PM. The cell-type specificity of these signatures was confirmed by single-cell RNA-sequencing data from the Human Protein Atlas^37^, where these signatures showed at least 13-fold and 110-fold enrichment in T-cells and B-cells, respectively, compared to other cell types (**Fig. 1a**). The T-cell score correlated with immunohistochemistry (IHC)-based CD4^+^ T-cell infiltration (Spearman r: 0.6301; p-value: 0.0029) and CD8^+^ T-cell infiltration (Spearman r: 0.7293; p-value: 0.0003) in the INITIATE^15^ cohort (n = 21), but not with PD-L1 expression (Spearman r: 0.2180; p-value: 0.3558) (**Fig. 1b-d**).

**Figure 1:**
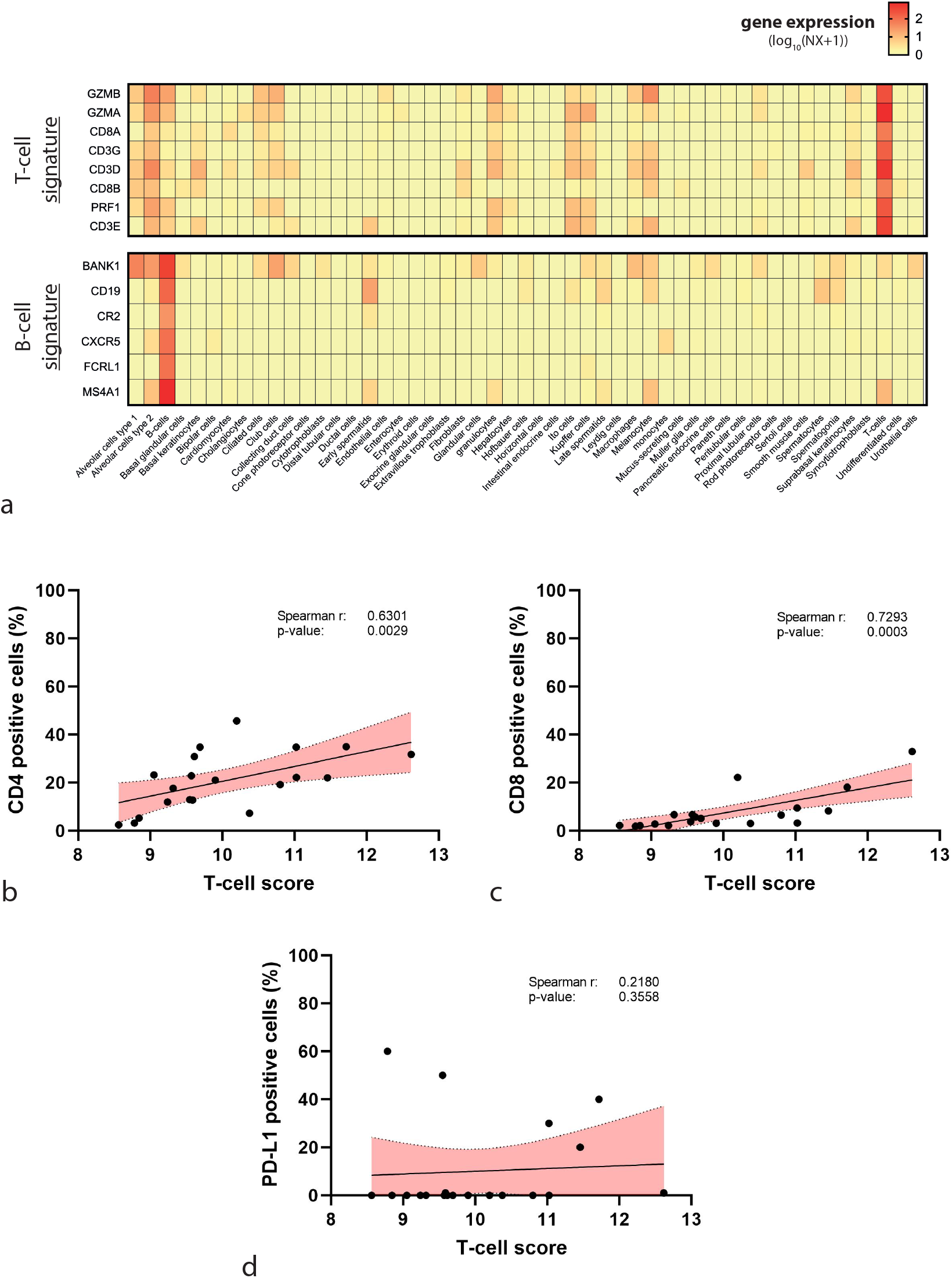
T- and B-cell transcriptional signatures. **a**) The T- and B-cell signature genes are specifically expressed in T-cells and B-cells, respectively, in the single-cell RNA-sequencing dataset from The Human Protein Atlas. (**b-d**) Correlation between sample T-cell score and IHC-based (**b**) CD4^+^ T-cells, (**c**) CD8^+^ T-cells, and (**d**) PD-L1 expression in samples from the INITIATE trial (n = 21). Red shading indicates the 95% confidence interval around a linear regression line.

### Immune profiling in ICB-naïve PM cohorts

Through unsupervised hierarchical clustering we identified three recurrent immune phenotypes in ICB-naïve PM patients, group one: T-cell ^--^;B-cell ^--^, group two: T-cell ^+^;B-cell ^++^, and group three: T-cell ^++^;B-cell ^+^, with an additional “rest” group for all other intermediate phenotypes (**Fig. 2a-c**). The frequency distribution of these groups was similar across cohorts, with 29.3-40.3% classified as T-cell ^--^;B-cell ^--^, 9.0-21.3% as T-cell ^+^;B-cell ^++^, and 24.0-31.8% as T-cell ^++^;B-cell ^+^ (**Fig. 2d**).

**Figure 2:**
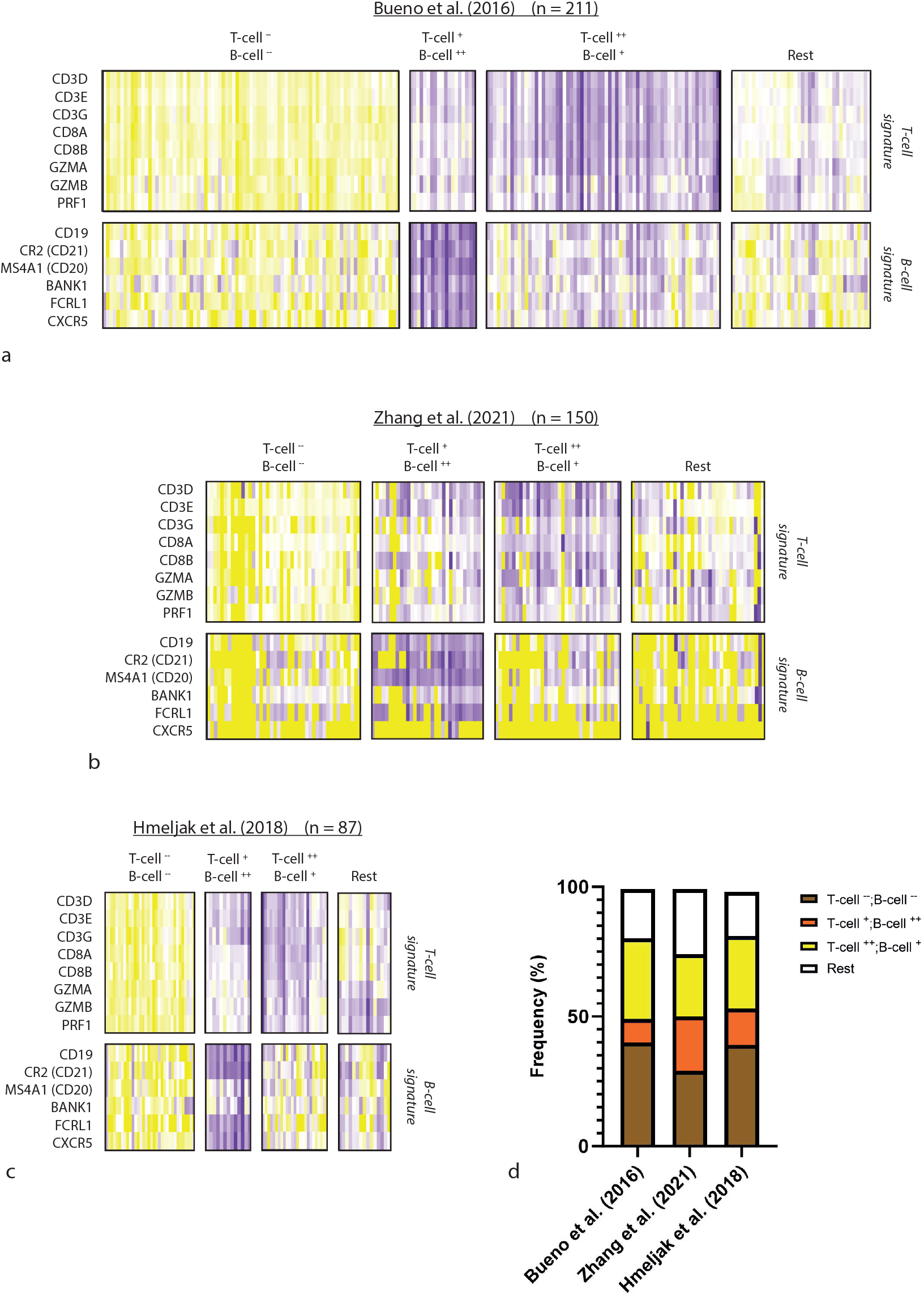
Landscape of T- and B-cell signature gene expression in three ICB-naïve PM cohorts: (**a**) Bueno et al. (2016), (**b**) Zhang et al. (2021), (**c**) Hmeljak et al. (2018). (**d**) Frequency of the recurrent immune subtypes in the three ICB-naïve PM cohorts.

Kaplan-Meier survival analysis was performed to evaluate the prognostic value of these immune subgroups in ICB-naïve PM patients, and we found that the B-cell-high subgroup (T-cell ^+^;B-cell ^++^) was associated with improved overall survival in the Bueno^21^, Zhang^29^ cohorts, but not in the Hmeljak^30^ cohort (**Fig. 3a-c**). Approximately 10-20% of PM patients have significantly increased B-cell score (**Fig. 2d**), and a threshold-based analysis of patients with the 10% highest B-cell scores confirmed its prognostic value in the Bueno^21^ (HR: 0.61, 95% CI: 0.39-0.94, p-value: 0.0269) and Zhang^29^ (HR: 0.50, 95% CI: 0.31-0.82, p-value: 0.0058) cohorts, but not the Hmeljak^30^ cohort (HR: 0.83, 95% CI: 0.37-1.846, p-value: 0.6452) (**Fig. 3d-f**).

**Figure 3:**
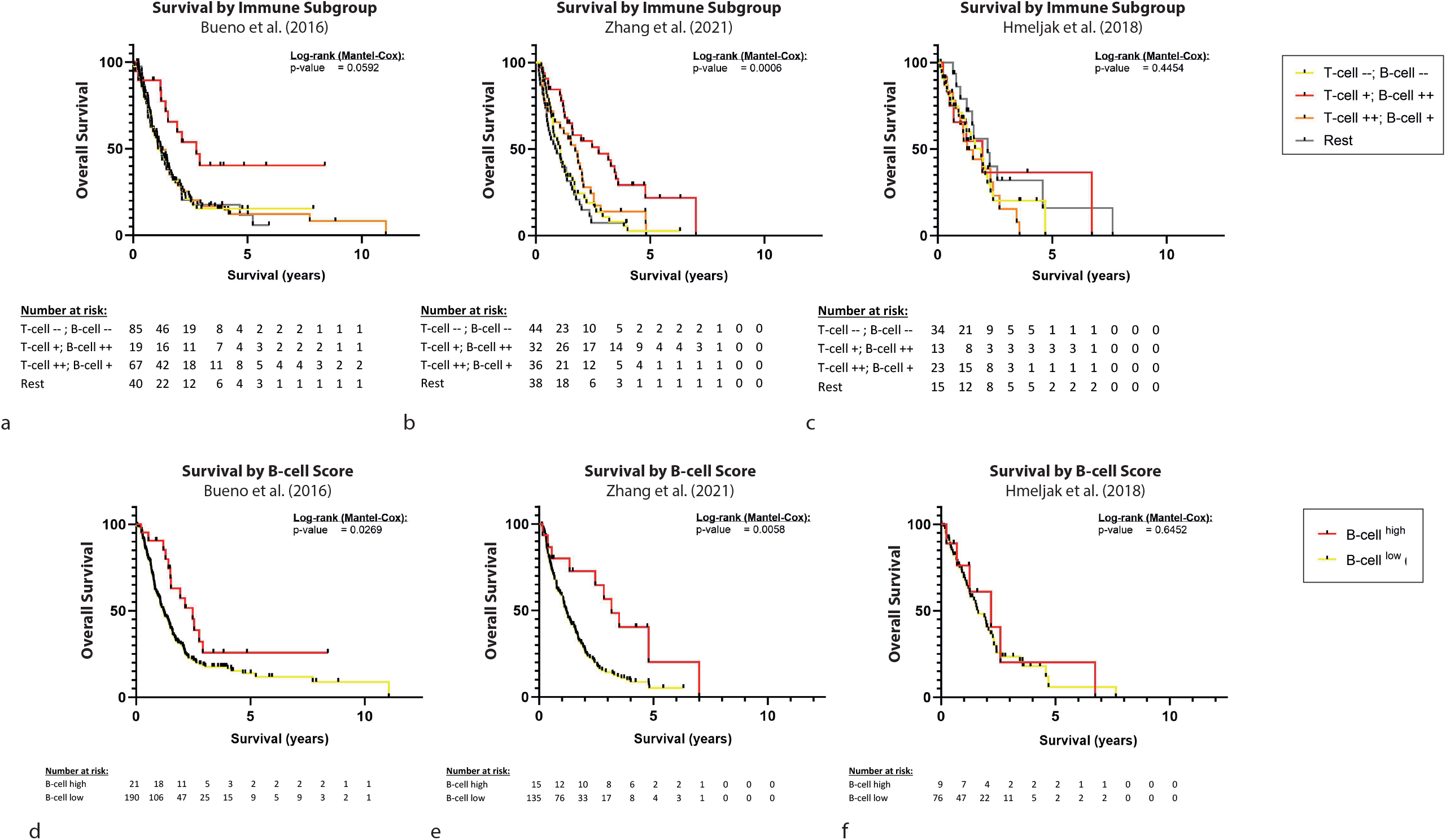
Survival of ICB-naive PM patients based on T- and B-cell immune infiltration. (**a-c**) Overall survival in the four recurrent T- and B-cell based immune subtypes in the three ICB-naïve PM cohorts. (**d-f**) Overall survival of the top 10% B-cell high versus bottom 90% B-cell low tumors in the three ICB-naïve PM cohorts.

To test for interaction between T- and B-cell score and histological subtype, these factors were included as covariates in a Cox regression analysis (**Suppl. Fig. 1**), demonstrating that the T-cell ^+^;B-cell ^++^ subgroup remained significantly associated with improved overall survival in both the Bueno^21^ (HR: 0.45, 95% CI: 0.21-0.84, p: 0.019) and Zhang^29^ (HR: 0.53, 95% CI: 0.32-0.85, p: 0.010) cohorts.

### Predictive value of T- and B-cells for ICB response

Next, we assessed the predictive value of T- and B-cell score in ICB-treated PM patients. An optimal cut-off threshold for T- and B-cell score was determined by testing quartile-based cut-off levels (corrected for multiple-testing) in the INITIATE^15^ (2^nd^ line nivolumab plus ipilimumab) and NivoMes^16^ (2^nd^ line nivolumab-only) cohorts. An optimal cut-off threshold for T-cell infiltration was determined at the top three quartiles (75%; high) vs. bottom quartile (25%; low), while B-cell infiltration was not significant at any cut-off level (**Suppl. Fig. 2**).

Expression of the T-cell score was significantly higher in responders to 2^nd^ line nivolumab plus ipilimumab (**Fig. 4a**), and high T-cell score predicted significantly improved mOS in these patients (HR: 0.06, 95% CI: 0.01-0.36, p-value: 0.0021) (**Fig. 4b**). This effect was not observed in the nivolumab-only group (HR: 0.84, 95% CI: 0.33-2.13, p-value: 0.6552) (**Fig. 4c,d**). In contrast, B-cell score had no predictive value for mOS across cut-off levels (**Suppl. Fig. 2**).

**Figure 4:**
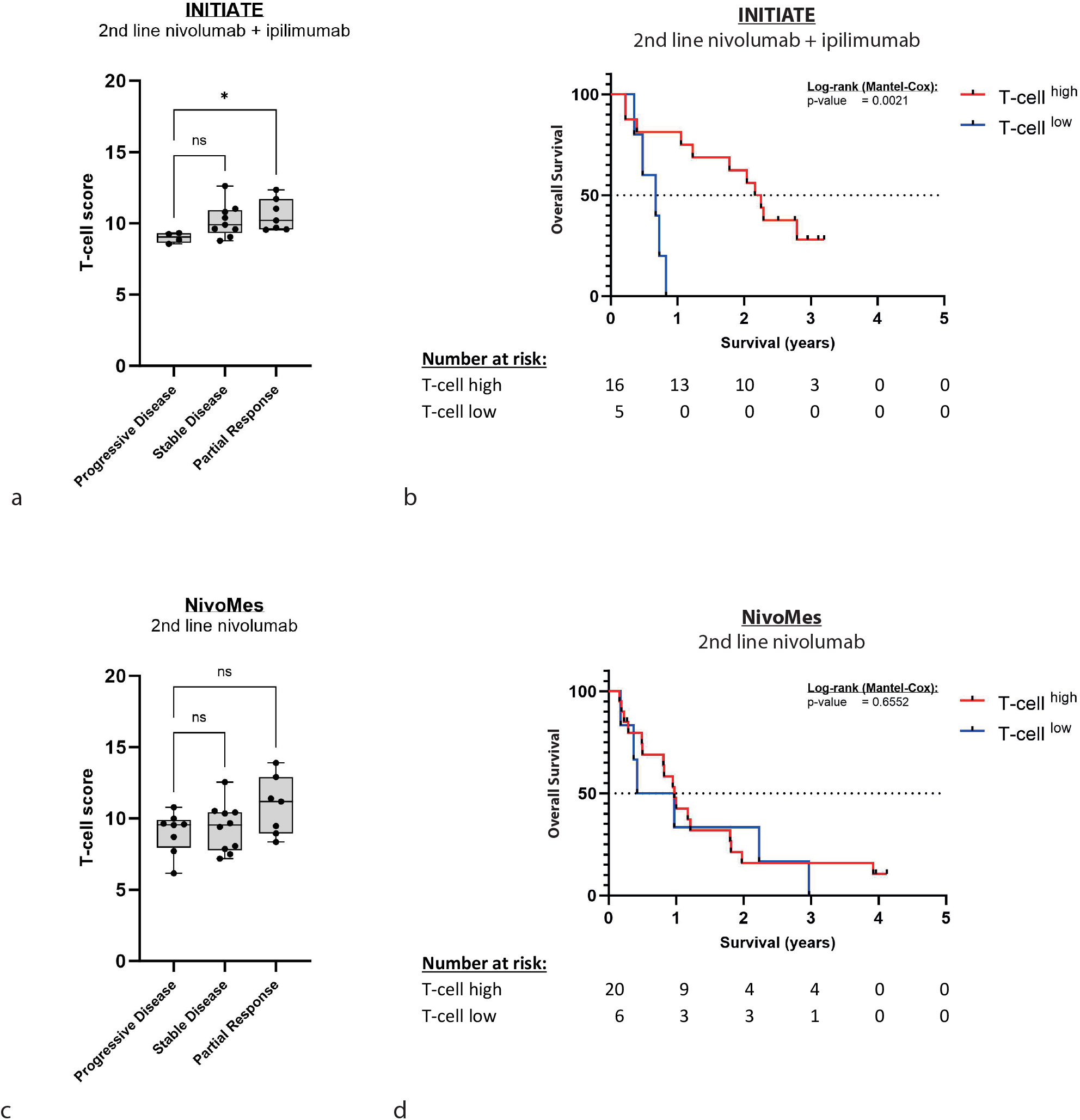
Association between T-cell score, objective response, and overall survival in PM patients. (**a**,**c**) Expression of T-cell score stratified by treatment response in the INITIATE and NivoMes cohorts. (**b**,**d**) Overall survival in T-cell low (bottom 25%) versus T-cell high (top 75%) PM tumors in the INITIATE, and NivoMes cohorts.

Expanding the mOS analysis to patients treated with an anti-PD1/PDL1 drug combined with non-immunotherapeutic drugs^31-33^ (**Fig. 5**), we found that patients with high T-cell score (top three quartiles; 75%) showed a trend towards improved survival after 2^nd^ line dostarlimab plus niraparib^33^ (HR: 0.26, 95% CI: 0.06-1.06). T-cell score had no predictive value for 2^nd^ line pembrolizumab plus bemcentinib^31^ (HR: 2.56, 95% CI: 0.66-9.86) or 2^nd^ line atezolizumab plus bevacizumab^32^ (HR: 0.46, 95% CI: 0.06-3.45). Importantly, the T-cell score had no overall prognostic value in ICB-naïve patients (**Fig. 5**), indicating the specificity of this marker for predicting response to 2^nd^ line nivolumab plus ipilimumab.

**Figure 5:**
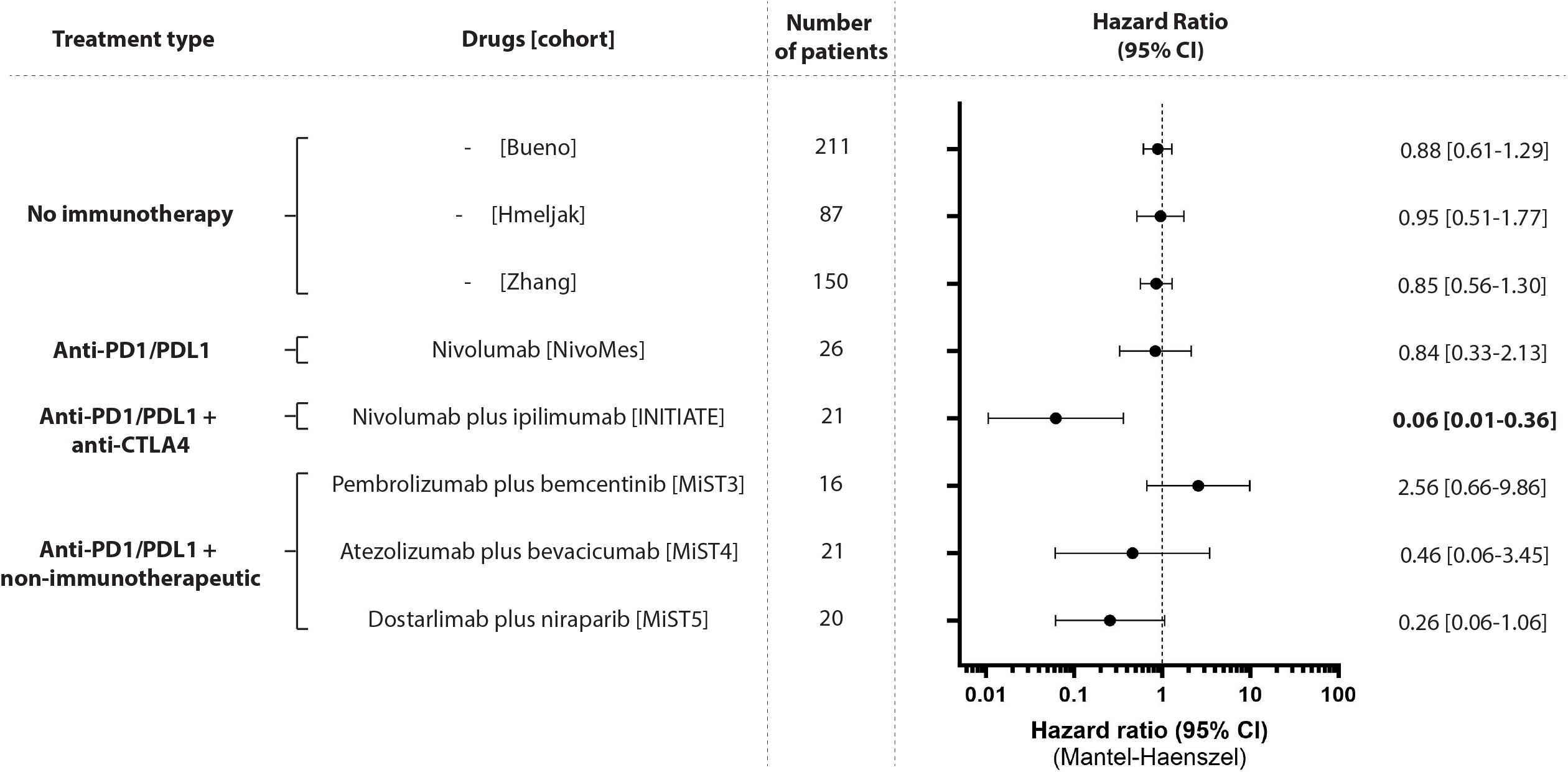
Hazard ratio for survival in the T-cell low (bottom 25%) versus T-cell high (top 75%) subgroups in the three ICB-naïve and five ICB-treated PM cohorts.

In summary, our eight-gene T-cell signature is a specific predictive biomarker for the combination of nivolumab (anti-PD1/PDL1) plus ipilimumab (anti-CTLA4) (**Fig. 4a,b**), but not for anti-PD1 monotherapy (**Fig. 4c,d**) or other non-immunotherapeutic drug combinations (**Fig. 5**).

## Discussion

This study explored the predictive and prognostic value of an eight-gene T-cell signature and a six-gene B-cell signature for ICB response in PM. Our results indicate that T-cell infiltration is a specific predictive biomarker for overall survival after combination treatment with 2^nd^ line nivolumab plus ipilimumab, but not for anti-PD1 monotherapy or combinations of anti-PD1/PDL1 with non-immunotherapeutic drugs. The T-cell signature did not show overall prognostic value in ICB-naïve patients, indicating its potential clinical utility as a specific marker for overall survival after treatment with 1^st^ or 2^nd^ line treatment with nivolumab plus ipilimumab in PM.

Clinical responses to 1^st^ and 2^nd^ line nivolumab plus ipilimumab in PM are variable^7,14-15^, highlighting the need for robust biomarkers. While PD-L1 expression has been widely investigated as a predictive biomarker for ICB in PM^7,9-16^, only two trials demonstrated significant predictive value among patients treated with nivolumab plus ipilimumab^14-15^. The INITIATE phase II trial of 2^nd^ line treatment with nivolumab plus ipilimumab in 36 patients found that PD-L1 expression (>1%) significantly predicted mOS (HR: 0.16, 95% CI: 0.04 - 0.73)^15^, while the non-comparative IFCT-1501 MAPS2 trial of 2^nd^ line treatment with nivolumab plus ipilimumab in 108 patients found that PD-L1 expression was associated with improved objective response rate, but not 12-week disease control rate or overall survival^14^. The larger phase III CheckMate 743 trial^7^, which compared 1^st^ line treatment with nivolumab plus ipilimumab to treatment with chemotherapy in 605 PM patients, found that while PD-L1 expression >1% predicted better mOS after treatment with nivolumab plus ipilimumab compared to chemotherapy (HR: 0.69, 95% CI: 0.55-0.87), it did not predict differences in mOS within the patient group who received nivolumab plus ipilimumab (17.3 months vs. 18.0 months for the <1% vs. ≥1% PD-L1 groups, respectively)^7^.

By establishing an RNA-based T-cell signature, we were able to retrospectively compare the prognostic and predictive value of T-cell infiltration in three large ICB-naïve^21,29-30^ (n = 448 patients) and five ICB-treated^15-16,31-33^ (n = 104 patients) PM cohorts. Our results indicate that this signature has potential for distinguishing between PM patients that do, or do not achieve clinical benefits after 2^nd^ line treatment with nivolumab plus ipilimumab. The signature did not predict clinical benefits for other anti-PD1/PDL1 treatment combinations, and did not have an overall prognostic value in ICB-naïve patients. These findings align with previous IHC-based analyses of CD4^+^ and CD8^+^ T-cell infiltration, which demonstrated that T-cell infiltration has predictive value for objective response rate after 2nd-line treatment with nivolumab plus ipilimumab^40^.

Infiltration of B-cells has been linked to improved outcomes after ICB treatment in other cancers^26-28^, but we did not find any significant association between B-cell signature expression and ICB response in PM. This inconsistency might in part be explained by the small proportion of PM patients with elevated B-cell infiltration (9.0-21.3%) combined with the small sample size of the clinical trial cohorts used in this study (16-26 patients). Larger clinical trial cohorts will be required to definitively clarify the predictive value of B-cell infiltration for ICB response in PM. Interestingly, we found that there is a subgroup of 9.0-21.3% of B-cell-high tumors among ICB-naïve patients, which is associated with a significantly improved mOS compared to B-cell-low tumors. This finding aligns with previous reports^41-43^ and suggests a role for B-cells in PM tumor control, potentially through an ongoing humoral anti-tumor response that is capable of suppressing remaining tumor cells after surgery.

Our study has several limitations. Due to the rare nature of PM and the small sample size of available phase II clinical trial cohorts (16-26 patients), our ability to evaluate the predictive value of these markers in the clinically relevant non-epithelioid subgroup is limited. The non-epithelioid subgroup was shown to derive the greatest benefit from 1^st^ line nivolumab plus ipilimumab^7^, and a larger randomized study in both epithelioid and non-epithelioid tumors will be required to assess the predictive value of the T-cell signature in this subgroup. In addition, PM tumors are physically large (median tumor volume between 100 and 620 cm^3^)^44^ and show significant genetic and micro-environmental intra-tumor heterogeneity^29,45^. Single-biopsy-based biomarkers may therefore not accurately capture whole-tumor heterogeneity. Finally, bulk RNA-sequencing data lacks the single-cell resolution required to identify more specific immune populations - such as exhausted T-cells - that might have an even stronger predictive value. Future studies based on spatial transcriptomics in ICB-treated PM patients will refine the predictive potential of these immune subpopulations.

In conclusion, we established high expression of an eight-gene T-cell signature as a potential biomarker for predicting response and mOS after 2^nd^ line nivolumab plus ipilimumab in PM. Larger randomized prospective studies in PM patients treated with 1^st^ line nivolumab plus ipilimumab are required to advance the clinical application of this biomarker.

## Data Availability

Most of the datasets analyzed in this study (Zhang et al., and the INITIATE, NivoMes, MiST3, MiST4, and MiST5 clinical trial datasets) are not publicly available due to patient privacy and data sharing restrictions. The authors do not have permission to redistribute these datasets. Processed data from the study by Hmeljak et al. (2018) were retrieved from Firebrowse (http://firebrowse.org/). Additional processed data and analysis scripts are available from the authors upon reasonable request.

## References

1. Selikoff, Irving J., Jacob Churg, and E. Cuyler Hammond. “Relation between exposure to asbestos and mesothelioma.” New England Journal of Medicine 272.11 (1965): 560–565.

2. Cancer Research UK: Survival for Mesothelioma. Available at https://www.cancerresearchuk.org/about-cancer/mesothelioma/survival. Accessed March 14, 2025.

3. American Cancer Society: Survival Rates for Mesothelioma. Available at https://www.cancer.org/cancer/types/malignant-mesothelioma/detection-diagnosis-staging/survival-statistics.html. Accessed March 14, 2025.

4. Vogelzang, Nicholas J., et al. “Phase III study of pemetrexed in combination with cisplatin versus cisplatin alone in patients with malignant pleural mesothelioma.” Journal of clinical oncology 21.14 (2003): 2636–2644.

5. Lim, Eric, et al. “Extended pleurectomy decortication and chemotherapy versus chemotherapy alone for pleural mesothelioma (MARS 2): a phase 3 randomised controlled trial.” The Lancet Respiratory Medicine 12.6 (2024): 457–466.

6. Ripley, R. Taylor, et al. “Going to MARS May Shorten Our Patient’s Survival.” The Journal of thoracic and cardiovascular surgery (2024): S0022–5223.

7. Baas, Paul, et al. “First-line nivolumab plus ipilimumab in unresectable malignant pleural mesothelioma (CheckMate 743): a multicentre, randomised, open-label, phase 3 trial.” The Lancet 397.10272 (2021): 375–386.

8. van Doorn, Sahar Barjesteh van Waalwijk, et al. “Adjuvant immune checkpoint blockade revisited.” The Lancet Oncology 24.7 (2023): 717–719.

9. Fennell, Dean A., et al. “Nivolumab versus placebo in patients with relapsed malignant mesothelioma (CONFIRM): a multicentre, double-blind, randomised, phase 3 trial.” The Lancet Oncology 22.11 (2021): 1530–1540.

10. Forde, Patrick M., et al. “Durvalumab with platinum-pemetrexed for unresectable pleural mesothelioma: survival, genomic and immunologic analyses from the phase 2 PrE0505 trial.” Nature medicine 27.11 (2021): 1910–1920.

11. Yap, Timothy A., et al. “Efficacy and safety of pembrolizumab in patients with advanced mesothelioma in the open-label, single-arm, phase 2 KEYNOTE-158 study.” The lancet respiratory medicine 9.6 (2021): 613–621.

12. Popat, S., et al. “A multicentre randomised phase III trial comparing pembrolizumab versus single-agent chemotherapy for advanced pre-treated malignant pleural mesothelioma: the European Thoracic Oncology Platform (ETOP 9-15) PROMISE-meso trial.” Annals of Oncology 31.12 (2020): 1734–1745.

13. Nowak, Anna K., et al. “Durvalumab with first-line chemotherapy in previously untreated malignant pleural mesothelioma (DREAM): a multicentre, single-arm, phase 2 trial with a safety run-in.” The Lancet Oncology 21.9 (2020): 1213–1223.

14. Scherpereel, Arnaud, et al. “Nivolumab or nivolumab plus ipilimumab in patients with relapsed malignant pleural mesothelioma (IFCT-1501 MAPS2): a multicentre, open label, randomised, non-comparative, phase 2 trial.” The Lancet Oncology 20.2 (2019): 239–253.

15. Disselhorst, Maria J., et al. “Ipilimumab and nivolumab in the treatment of recurrent malignant pleural mesothelioma (INITIATE): results of a prospective, single-arm, phase 2 trial.” The Lancet Respiratory Medicine 7.3 (2019): 260–270.

16. Quispel-Janssen, Josine, et al. “Programmed death 1 blockade with nivolumab in patients with recurrent malignant pleural mesothelioma.” Journal of Thoracic Oncology 13.10 (2018): 1569–1576.

17. Pilard, Charlotte, et al. “Cancer immunotherapy: it’s time to better predict patients’ response.” British journal of cancer 125.7 (2021): 927–938.

18. Mansfield, Aaron S., et al. “Neoantigenic potential of complex chromosomal rearrangements in mesothelioma.” Journal of Thoracic Oncology 14.2 (2019): 276–287.

19. Litchfield, Kevin, et al. “Meta-analysis of tumor-and T cell-intrinsic mechanisms of sensitization to checkpoint inhibition.” Cell 184.3 (2021): 596–614.

20. Budczies, Jan, et al. “Tumour mutational burden: clinical utility, challenges and emerging improvements.” Nature Reviews Clinical Oncology (2024): 1–18.

21. Bueno, Raphael, et al. “Comprehensive genomic analysis of malignant pleural mesothelioma identifies recurrent mutations, gene fusions and splicing alterations.” Nature genetics 48.4 (2016): 407–416.

22. Désage, Anne-Laure, et al. “The immune microenvironment of malignant pleural mesothelioma: A literature review.” Cancers 13.13 (2021):3205.

23. Hiltbrunner, Stefanie, et al. “Tumor immune microenvironment and genetic alterations in mesothelioma.” Frontiers in oncology 11 (2021): 660039.

24. Van der Leun, Anne M., Daniela S. Thommen, and Ton N. Schumacher. “CD8+ T cell states in human cancer: insights from single-cell analysis.” Nature Reviews Cancer 20.4 (2020): 218–232.

25. Budimir, Natalija, et al. “Reversing T-cell exhaustion in cancer: lessons learned from PD-1/PD-L1 immune checkpoint blockade.” Cancer immunology research 10.2 (2022): 146–153.

26. Helmink, Beth A., et al. “B cells and tertiary lymphoid structures promote immunotherapy response.” Nature 577.7791 (2020): 549–555.

27. Petitprez, Florent, et al. “B cells are associated with survival and immunotherapy response in sarcoma.” Nature 577.7791 (2020): 556–560.

28. Cabrita, Rita, et al. “Tertiary lymphoid structures improve immunotherapy and survival in melanoma.” Nature 577.7791 (2020): 561–565.

29. Zhang, Min, et al. “Clonal architecture in mesothelioma is prognostic and shapes the tumour microenvironment.” Nature communications 12.1 (2021): 1751.

30. Hmeljak, Julija, et al. “Integrative molecular characterization of malignant pleural mesothelioma.” Cancer discovery 8.12 (2018): 1548–1565.

31. MiST3 clinical trial [manuscript in preparation].

32. Zhang, Min, et al. “A gut microbiota rheostat forecasts responsiveness to PD-L1 and VEGF blockade in mesothelioma.” Nature Communications 15.1 (2024):7187.

33. MiST5 clinical trial [manuscript in preparation].

34. Love, Michael I., Wolfgang Huber, and Simon Anders. “Moderated estimation of fold change and dispersion for RNA-seq data with DESeq2.” Genome biology 15 (2014): 1 21.

35. Andreatta, Massimo, et al. “Interpretation of T cell states from single-cell transcriptomics data using reference atlases.” Nature communications 12.1 (2021):2965.

36. Playoust, Eve, et al. “Germinal center-dependent and-independent immune responses of tumor-infiltrating B cells in human cancers.” Cellular & Molecular Immunology 20.9 (2023): 1040–1050.

37. Uhlen, Mathias, et al. “Towards a knowledge-based human protein atlas.” Nature biotechnology 28.12 (2010): 1248–1250.

38. Vandesompele, Jo, et al. “Accurate normalization of real-time quantitative RT-PCR data by geometric averaging of multiple internal control genes.” Genome biology 3 (2002): 1–12.

39. Wilkerson, Matthew D., and D. Neil Hayes. “ConsensusClusterPlus: a class discovery tool with confidence assessments and item tracking.” Bioinformatics 26.12 (2010): 1572–1573.

40. Disselhorst, Maria J., et al. “Immune cells in mesothelioma microenvironment simplistic marker of response to nivolumab plus ipilimumab?.” Lung Cancer 173 (2022): 49–52.

41. Pasello, G., et al. “Malignant pleural mesothelioma immune microenvironment and checkpoint expression: correlation with clinical–pathological features and intratumor heterogeneity over time.” Annals of Oncology 29.5 (2018): 1258–1265.

42. Chee, Serena J., et al. “Evaluating the effect of immune cells on the outcome of patients with mesothelioma.” British journal of cancer 117.9 (2017): 1341–1348.

43. Ujiie, Hideki, et al. “The tumoral and stromal immune microenvironment in malignant pleural mesothelioma: a comprehensive analysis reveals prognostic immune markers.” Oncoimmunology 4.6 (2015): e1009285.

44. Murphy, David J., and Ritu R. Gill. “Volumetric assessment in malignant pleural mesothelioma.” Annals of translational medicine 5.11 (2017).

45. Meiller, Clément, et al. “Multi-site tumor sampling highlights molecular intra-tumor heterogeneity in malignant pleural mesothelioma.” Genome Medicine 13 (2021): 1–16

