## Supplementary Figure 1 for "The Predictive and Prognostic Value of T- and B-cell Transcriptomic Signatures for Clinical Response to Immune Checkpoint Blockade in Pleural Mesothelioma"

| Variable | | Number of samples | HR | 95% CI | p-value |
|---|---|---|---|---|---|
| Immune subgroup | T-cell -- ; B-cell -- | 85 | *Reference* | | |
| | T-cell +; B-cell ++ | 19 | 0.45 | 0.21-0.84 | 0.019 (*) |
| | T-cell ++; B-cell + | 67 | 0.93 | 0.65-1.38 | 0.717 |
| | Rest | 40 | 1.07 | 0.69-1.62 | 0.768 |
| Histology | Epithelioid | 141 | *Reference* | | |
| | Biphasic | 62 | 1.47 | 1.03-2.06 | 0.029 (*) |
| | Sarcomatoid | 7 | 2.02 | 0.78-4.29 | 0.100 |
| | Desmoplastic | 1 | 25.04 | 1.35-135.9 | 0.0024 (**) |

a

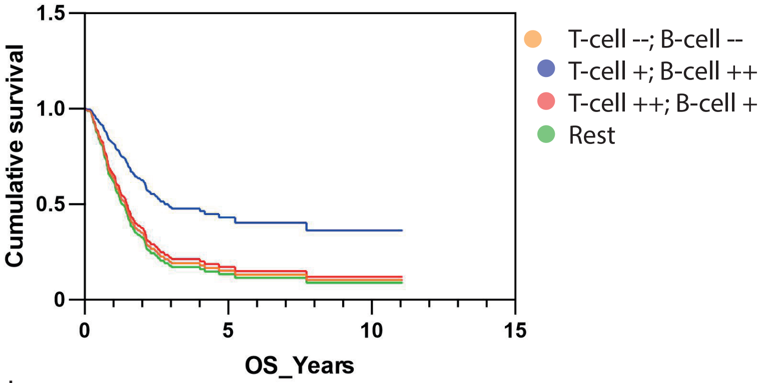


b

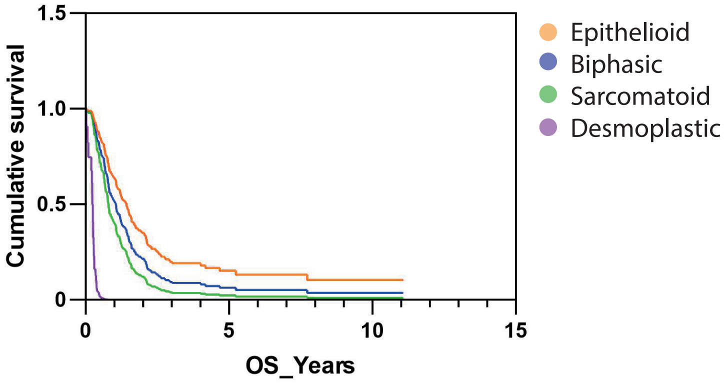


c

| Variable | | Number of samples | HR | 95% CI | p-value |
|---|---|---|---|---|---|
| Immune subgroup | T-cell -- ; B-cell -- | 44 | *Reference* | | |
| | T-cell +; B-cell ++ | 32 | 0.53 | 0.32-0.85 | 0.010 (*) |
| | T-cell ++; B-cell + | 36 | 1.00 | 0.66-1.50 | 0.986 |
| | Rest | 38 | 1.41 | 0.96-2.07 | 0.076 |
| Histology | Epithelioid | 137 | *Reference* | | |
| | Biphasic | 10 | 1.37 | 0.87-2.09 | 0.154 |
| | Sarcomatoid | 3 | 1.71 | 0.66-3.62 | 0.210 |

d

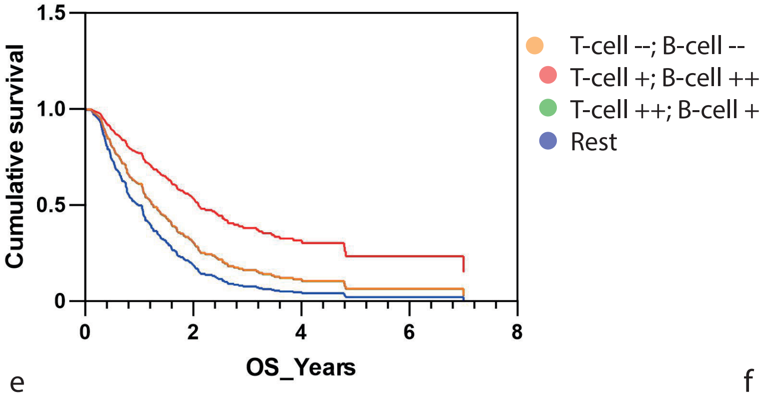


e

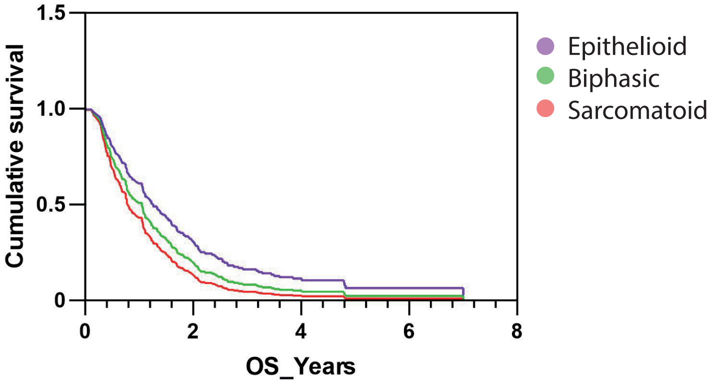


f

Supplementary Figure 1: Cox regression for the (a-c) Bueno et al. (2016) cohort, and the (d-e) Zhang et al. (2021) cohort.
