## Supplementary Figure 2 for "The Predictive and Prognostic Value of T- and B-cell Transcriptomic Signatures for Clinical Response to Immune Checkpoint Blockade in Pleural Mesothelioma"

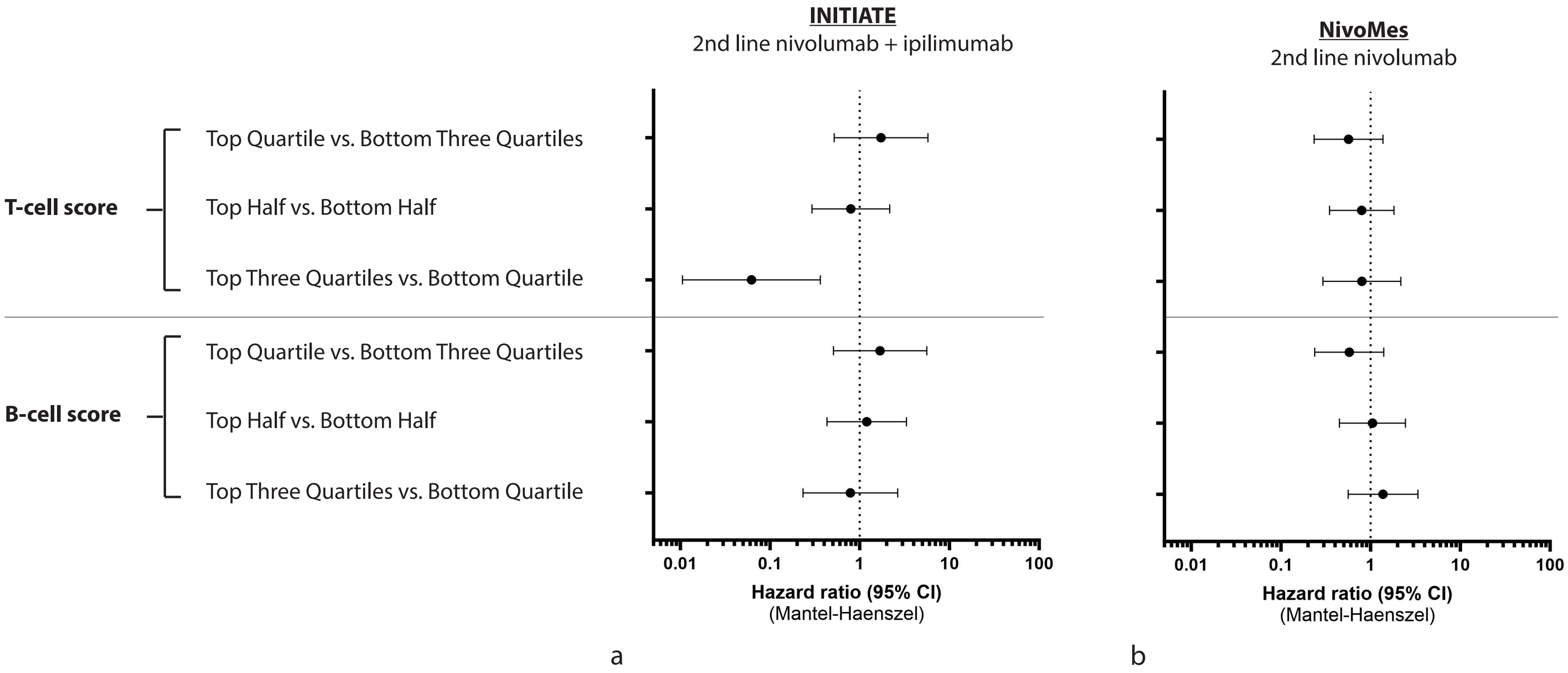


Supplementary Figure 2: Predictive value of T- and B-cell signatures in the (a) INITIATE and (b) NivoMes trials at different cut-of thresholds.
